# Cohort Profile Update: The HUNT Study, Norway

**DOI:** 10.1101/2021.10.12.21264858

**Authors:** Bjørn Olav Åsvold, Arnulf Langhammer, Tommy Aune Rehn, Grete Kjelvik, Trond Viggo Grøntvedt, Elin Pettersen Sørgjerd, Jørn Søberg Fenstad, Oddgeir Holmen, Maria C Stuifbergen, Sigrid Anna Aalberg Vikjord, Ben M Brumpton, Håvard Kjesbu Skjellegrind, Pernille Thingstad, Erik R Sund, Geir Selbæk, Paul Jarle Mork, Vegar Rangul, Kristian Hveem, Marit Næss, Steinar Krokstad

**Author notes:** Corresponding author: Bjørn Olav Åsvold, Department of Public Health and Nursing, NTNU, Norwegian University of Science and Technology, Postboks 8905 MTFS, NO-7491 Trondheim, Norway.

## Abstract

In the HUNT Study, all residents aged ≥20 years in the Nord-Trøndelag region, Norway have been invited to repeated surveys since 1984-86. The study data may be linked to local and national health registries. The HUNT4 survey in 2017-19 included 56 042 participants in Nord-Trøndelag and 107 711 participants in the neighboring Sør-Trøndelag region. The HUNT4 data enable more long-term follow-up, studies of life-course health trajectories, and within-family studies. New measures include body composition analysis using bioelectrical impedance; a one-week accelerometer recording; physical and cognitive testing in older adults; measurements of hemoglobin and blood cell counts, HbA1c and phosphatidylethanol; and genotyping. Researchers can apply for HUNT data access from HUNT Research Centre if they have obtained project approval from the Regional Committee for Medical and Health Research Ethics, see www.ntnu.edu/hunt/data.

## The original cohort

The Trøndelag Health Study (the HUNT Study) is a population-based cohort study of the adult population in Trøndelag County, Norway. It was previously called the Nord-Trøndelag Health Study, until the study in 2019 expanded to cover both regions of Trøndelag County, Nord-Trøndelag and Sør-Trøndelag. The study has been running in Nord-Trøndelag since 1984 and is designed to cover a broad range of health-related topics through repeated surveys with questionnaires, interviews, clinical examinations, laboratory measurements and storage of biological samples. Nord-Trøndelag is fairly representative of Norway except for the lack of large cities and immigrant populations, and the region is suitable for longitudinal studies due to low migration. The HUNT Study data can be linked to a wide range of local and national health registries by means of the unique identification number allocated to all Norwegian residents. All residents ≥20 years of age in Nord-Trøndelag were invited to the HUNT1 (1984-86, 77 202 participants, 89.4% of invitees participated),^1^ HUNT2 (1995-97, 65 228 participants, 69.5%)^2^ and HUNT3 (2006-08, 50 800 participants, 54.1%)^3^ surveys (Figure 1). Since 1995-97, all adolescents (13-19 years of age) in Nord-Trøndelag have been invited to participate in the corresponding Young-HUNT Study.^4^

**Figure 1.**
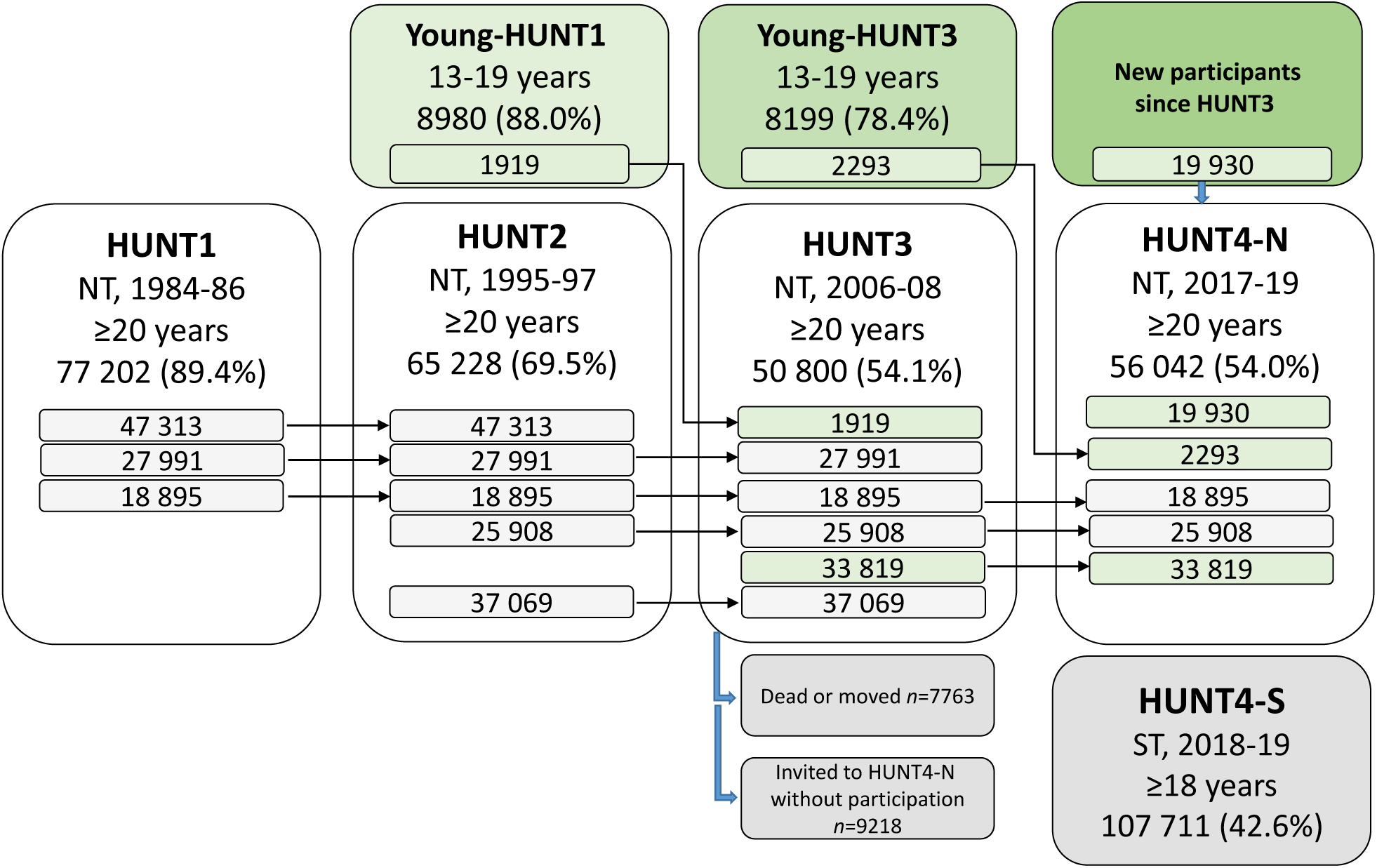
Flowchart of participation across the HUNT1-HUNT4 surveys and details on recruitment to HUNT4. The few individuals who have withdrawn their consent to participate in the HUNT Study have been excluded, and the numbers therefore deviate slightly from those reported in previous HUNT cohort profiles. NT, Nord-Trøndelag region; ST, Sør-Trøndelag region.

### What is the reason for the new data collection?

In 2017-19, all adult residents of Trøndelag were invited to the HUNT4 survey. The reasons for this new survey were first, to collect follow-up and extended information on previous participants, and new information on individuals having moved into the region or reached adult age. After completion of HUNT4, about 19 000 individuals have more than 30 years of follow-up spanning all four study cycles (Figure 1). The continued follow-up also allows for monitoring of secular trends in public health. Second, improved measurement methods have become available for several health and lifestyle characteristics important for public health, as described in the later sections. Third, the HUNT Study previously lacked data from a large city. In 2019, HUNT4 expanded to include a survey in Sør-Trøndelag, which includes the city of Trondheim with ∼202 000 inhabitants (2019). The extended study population from both Nord-Trøndelag and Sør-Trøndelag is generally representative of Norway (Supplementary Table S1). A fourth reason for new data collection is the expansion of genetic and molecular epidemiology, where 70 517 participants of the HUNT2 or HUNT3 surveys have been genotyped.

### What will be the new areas of research?

Extended follow-up of HUNT participants and high attendance among older adults enable a stronger focus on disease incidence, progression and life-course health trajectories. New generations of HUNT participants enable more within-family and inter-generational studies. More valid studies of physical activity and anthropometry are possible due to new data including accelerometer recordings and body composition analyses using bioelectrical impedance. Other new assessments include cognitive and physical testing in old age, fecal sampling for microbiome studies, and metabolomics and proteomics analyses for deeper molecular phenotyping. With available genotype information, analyses are expanded to e.g. genome-wide association studies and Mendelian randomization.

### Who is in the cohort?

In Nord-Trøndelag, all residents aged ≥20 years at the estimated time of survey participation were invited to HUNT4 between 29 August 2017 and 23 February 2019 (the HUNT4-N Survey). Out of 103 800 invitees, 56 042 (54.0%) participated, defined as returning the main questionnaire.Participation across the HUNT1-HUNT4 surveys is shown in Figure 1. In Sør-Trøndelag, all residents aged ≥18 years were invited to the HUNT4-S Survey and 107 711 (42.6%) participated out of 252 991 invitees. HUNT4-S consisted of two parts: 1745 residents ≥70 years of age in districts of Trondheim municipality took part in an examination between 26 Oct 2018 and 12 Jun 2019, and the remaining took part in a questionnaire survey between 3 Oct 2019 and 19 Nov 2019.

Characteristics of HUNT4 participants are given in Table 1. Participation in HUNT4 differed by age, being highest in the age groups 40-79 years, and also slightly differed by marital status and urban vs. rural residency (Table 2). Attrition from HUNT3 to HUNT4 was highest in older adults and was also moderately higher among people with chronic diseases, poor self-rated health, or who were smokers (Supplementary Table S2). Questionnaire surveys were performed among nonparticipants of HUNT4-N and previous HUNT participants who were not invited to HUNT4-N because they had emigrated from the study area (Table 3, Supplementary Table S3). Those who had emigrated reported lower BMI, more physical activity, less smoking and better self-rated health and more frequent alcohol intake than both participants and nonparticipants. The nonparticipants had less healthy lifestyle and lower self-reported health and higher proportion of cardiovascular diseases, chronic obstructive pulmonary disease, diabetes and antihypertensive medication use.

**Table 1.** Characteristics of participants in the HUNT4 Survey (HUNT4-N and HUNT4-S), reported as percentages^a^ unless otherwise noted.

| Characteristics | HUNT4-N<sup>b</sup> |  | HUNT4-S<sup>c</sup> |  |
| --- | --- | --- | --- | --- |
|  | Women<br>(n=30 574) | Men<br>(n=25 468) | Women<br>(n=61 066) | Men<br>(n=46 645) |
| Age (years), mean (SD, range) | 53.9 (17.8,<br>19-102)<sup>d</sup> | 54.8 (17.3,<br>19-101)<sup>d</sup> | 49.9 (17.9,<br>18-105) | 52.2 (17.7,<br>18-100) |
| Age groups (years) |  |  |  |  |
| <30 | 11.9 | 11.0 | 16.8 | 13.7 |
| 30-39 | 12.7 | 11.3 | 15.7 | 13.6 |
| 40-49 | 16.6 | 15.4 | 17.2 | 16.7 |
| 50-59 | 19.0 | 19.4 | 19.2 | 19.4 |
| 60-69 | 18.8 | 21.3 | 16.1 | 18.6 |
| 70-79 | 14.2 | 15.6 | 10.2 | 13.2 |
| 80-89 | 5.6 | 5.2 | 3.9 | 4.4 |
| ≥90 | 1.2 | 0.7 | 0.8 | 0.5 |
| Years of education<sup>e</sup> |  |  |  |  |
| ≤10 | 13.1 | 10.0 | 7.9 | 6.5 |
| 11-13 | 44.4 | 55.3 | 39.2 | 44.2 |
| ≥14 | 42.5 | 34.7 | 52.9 | 49.3 |
| Married, registered partner or cohabitant | 71.5 | 77.4 | 67.5 | 72.7 |
| Living in urban constituency | 39.6 | 37.3 | 56.3 | 56.3 |
| Good or very good self-rated health | 73.6 | 78.6 | 74.2 | 80.5 |
| Daily smoking |  |  |  |  |
| Never | 56.1 | 55.9 | 71.2 | 68.9 |
| Former | 34.3 | 37.1 | 21.3 | 25.2 |
| Current | 9.6 | 7.0 | 7.5 | 5.9 |
| Alcohol, units per week (median, IQR)<sup>f</sup> | 1 (0-2.5) | 2.5 (1-4.5) | 1 (0-3) | 3 (1-5.5) |
| Frequency of physical exercise |  |  |  |  |
| ≤ once/week | 32.4 | 40.6 | 28.7 | 34.0 |
| 2-3 times/week | 43.9 | 39.0 | 42.4 | 40.3 |
| Almost every day | 23.7 | 20.4 | 29.0 | 25.7 |
| Self-reported history of current or previous disease |  |  |  |  |
| Coronary heart disease (myocardial infarction or angina pectoris) | 3.5 | 8.6 | 3.2 | 8.3 |
| Atrial fibrillation | 4.0 | 7.0 | 4.3 | 7.9 |
| Stroke | 2.9 | 4.1 | 2.1 | 3.3 |
| Asthma | 12.5 | 11.6 | 14.4 | 13.1 |
| COPD | 2.6 | 3.3 | 1.9 | 2.6 |
| Diabetes | 5.2 | 7.1 | 3.9 | 6.1 |
| Hypo- or hyperthyroidism | 12.1 | 3.3 | 12.1 | 3.7 |
| Cancer | 7.9 | 8.0 | 7.8 | 8.1 |
| Sought health care for mental health problems | 21.6 | 12.2 | 32.9 | 19.5 |
| Current drug treatment |  |  |  |  |
| Antihypertensive medication | 21.4 | 25.6 | 15.6 | 21.8 |
| Lipid-lowering medication | 14.3 | 18.8 | 9.9 | 17.2 |
| Medication for asthma or COPD | 8.3 | 7.3 | 7.5 | 6.6 |
| Medication for anxiety or depression | 8.9 | 4.7 | 6.6 | 3.9 |
| Medication for thyroid dysfunction | 9.5 | 2.7 | 8.8 | 2.6 |
| Visit to a general practitioner during the last 12 months | 85.9 | 77.7 | 86.2 | 78.1 |
| Admitted to hospital during the last 12 months | 14.1 | 12.4 | NA | NA |
| Body mass index<sup>g</sup> (kg/m<sup>2</sup>), mean (SD) | 26.9 (5.1) | 27.6 (4.2) | 25.8 (4.9) | 26.7 (4.1) |
| Body mass index categories<sup>g</sup> (kg/m<sup>2</sup>) |  |  |  |  |
| <18.5 | 1.5 | 0.4 | 1.7 | 0.5 |
| 18.5-24.9 | 38.2 | 27.1 | 48.6 | 36.7 |
| 25.0-29.9 | 36.3 | 48.3 | 32.6 | 45.7 |
| ≥30.0 | 24.1 | 24.2 | 17.1 | 17.1 |
| Systolic blood pressure<sup>h</sup> (mmHg), mean (SD) | 126.1 (19.4) | 131.0 (16.9) | NA | NA |
| Diastolic blood pressure<sup>h</sup> (mmHg), mean (SD) | 70.8 (9.4) | 76.0 (10.3) | NA | NA |
| Low-density lipoprotein cholesterol<sup>i</sup> (mmol/l), mean (SD) | 3.25 (0.97) | 3.15 (0.96) | NA | NA |
| Estimated glomerular filtration rate<sup>j</sup> <60 ml/min/1.73m<sup>2</sup> | 6.6 | 6.2 | NA | NA |

Abbreviations: COPD, chronic obstructive pulmonary disease; IQR, inter-quartile range; SD, standard deviation
<sup>a</sup> Calculated among people with valid information, which ranges from 89% to 100% for individual items
<sup>b</sup> The survey was conducted in Nord-Trøndelag between 29 August 2017 and 23 February 2019 (the survey includes a pilot study performed in May 2017). Characteristics are presented for the 56 042 individuals who participated in HUNT4-N by returning the main questionnaire. In addition, 897 individuals took part in the clinical examination, but did not return the questionnaire; these individuals were more often older adults included in the HUNT70+ study part that will be described in a separate paper.
<sup>c</sup> The survey was conducted in Sør-Trøndelag as a questionnaire survey between 3 October and 19 November 2019 (105 966 participants) or as an examination among residents ≥70 years in districts of Trondheim municipality between 26 October 2018 and 12 June 2019 (1745 participants). The 252 991 invitees to HUNT4-S include all adult residents of Sør-Trøndelag, except former Nord-Trøndelag residents who had moved to Sør-Trøndelag after previously participating in HUNT and were instead invited to an emigrant survey.
<sup>d</sup> All residents aged ≥20 years at the estimated time of survey participation were invited, and some were aged 19 years at the date of participation.
<sup>e</sup> Years of education based on self-reported level of education: primary school ≤10 years; academic or vocational school/apprentice 11-13 years; and university college or university ≥14 years.
<sup>f</sup> Units (12.8 g alcohol) calculated based on the self-reported usual number of glasses of beer/wine/liquor per 2 weeks or, if that information was missing, estimated from the reported frequency of alcohol consumption during the last year.
<sup>g</sup> For participants of the questionnaire survey in HUNT4-S, this was calculated from self-reported weight and height
<sup>h</sup> Mean of the 2<sup>nd</sup> and 3<sup>rd</sup> measurements. For 0.4% of participants who had only 2 valid blood pressure measurements, we used the 2<sup>nd</sup> measurement.
<sup>i</sup> Estimated using the equation developed by Sampson M et al., *JAMA Cardiol.* 2020;5(5):540-548.
<sup>j</sup> Estimated using the Chronic Kidney Disease Epidemiology collaboration (CKD-EPI) equation

**Table 2.** Relative probability of participation in HUNT4-N and HUNT4-S by sociodemographic characteristics.

| Characteristics | HUNT4-N |  |  |  | HUNT4-S |  |  |  |
| --- | --- | --- | --- | --- | --- | --- | --- | --- |
|  | Women |  | Men |  | Women |  | Men |  |
|  | Participants,<br>*n* (%)<sup>a</sup> | Relative<br>probability<sup>b</sup><br>(95% CI) | Participants,<br>*n* (%)<sup>a</sup> | Relative<br>probability<sup>b</sup><br>(95% CI) | Participants,<br>*n* (%)<sup>a</sup> | Relative<br>probability<sup>b</sup><br>(95% CI) | Participants,<br>*n* (%)<sup>a</sup> | Relative<br>probability<sup>b</sup><br>(95% CI) |
| Total invited population | 30 574<br>(58.8%) |  | 25 468<br>(49.1%) |  | 61 066<br>(48.9%) |  | 46 645<br>(36.4%) |  |
| Age (years) |  |  |  |  |  |  |  |  |
| <30 | 3643<br>(43.5) | 1.00<br>(reference) | 2811<br>(31.0) | 1.00<br>(reference) | 10 289<br>(38.4) | 1.00<br>(reference) | 6396<br>(21.6) | 1.00<br>(reference) |
| 30-39 | 3870<br>(52.5) | 1.21<br>(1.17-1.25) | 2881<br>(38.1) | 1.23<br>(1.18-1.28) | 9591<br>(46.3) | 1.20<br>(1.18-1.23) | 6327<br>(27.9) | 1.29<br>(1.26-1.33) |
| 40-49 | 5087<br>(59.1) | 1.36<br>(1.32-1.40) | 3908<br>(45.2) | 1.46<br>(1.40-1.52) | 10 532<br>(52.9) | 1.38<br>(1.35-1.40) | 7784<br>(36.8) | 1.70<br>(1.66-1.75) |
| 50-59 | 5817<br>(65.7) | 1.51<br>(1.47-1.56) | 4948<br>(53.9) | 1.74<br>(1.68-1.80) | 11 728<br>(59.5) | 1.55<br>(1.52-1.58) | 9044<br>(44.4) | 2.06<br>(2.00-2.11) |
| 60-69 | 5745<br>(69.9) | 1.61<br>(1.56-1.65) | 5430<br>(63.8) | 2.06<br>(1.99-2.13) | 9851<br>(59.3) | 1.54<br>(1.51-1.57) | 8677<br>(52.1) | 2.41<br>(2.35-2.48) |
| 70-79 | 4329<br>(67.8) | 1.56<br>(1.51-1.61) | 3978<br>(65.3) | 2.11<br>(2.03-2.18) | 6253<br>(47.5) | 1.24<br>(1.21-1.26) | 6151<br>(49.5) | 2.29<br>(2.23-2.36) |
| 80-89 | 1725<br>(54.1) | 1.25<br>(1.20-1.30) | 1327<br>(56.5) | 1.82<br>(1.74-1.91) | 2357<br>(38.3) | 1.00<br>(0.96-1.03) | 2041<br>(46.0) | 2.13<br>(2.05-2.21) |
| ≥90 | 358<br>(37.5) | 0.86<br>(0.79-0.94) | 185<br>(41.9) | 1.35<br>(1.20-1.51) | 465<br>(25.2) | 0.66<br>(0.61-0.71) | 225<br>(29.7) | 1.38<br>(1.23-1.54) |
| Marital status |  |  |  |  |  |  |  |  |
| Married or registered partner | 15 184<br>(66.5) | 1.00<br>(reference) | 14 092<br>(60.3) | 1.00<br>(reference) | 27 230<br>(55.7) | 1.00<br>(reference) | 24 033<br>(47.2) | 1.00<br>(reference) |
| Not married or<br>registered partner | 8775<br>(50.2) | 0.89<br>(0.87-0.90) | 8130<br>(37.2) | 0.78<br>(0.76-0.80) | 22 687<br>(44.6) | 0.92<br>(0.91-0.93) | 16 848<br>(26.9) | 0.76<br>(0.74-0.77) |
| Divorced,<br>separated or<br>widow(er) | 6549<br>(56.6) | 0.86<br>(0.85-0.88) | 3188<br>(48.5) | 0.79<br>(0.76-0.81) | 11 085<br>(44.2) | 0.83<br>(0.82-0.85) | 5743<br>(39.8) | 0.81<br>(0.79-0.82) |
| Constituency type |  |  |  |  |  |  |  |  |
| Urban | 12 063<br>(55.5) | 1.00<br>(reference) | 9470<br>(46.4) | 1.00<br>(reference) | 33 399<br>(46.2) | 1.00<br>(reference) | 25 537<br>(34.3) | 1.00<br>(reference) |
| Rural | 18 411<br>(61.1) | 1.09<br>(1.07-1.11) | 15 921<br>(50.8) | 1.07<br>(1.05-1.09) | 26 660<br>(53.6) | 1.14<br>(1.13-1.15) | 20 371<br>(39.7) | 1.04<br>(1.03-1.06) |

<sup>a</sup> Percentage among the total number of invitees in the category
<sup>b</sup> Age-adjusted relative probability of participation

**Table 3.**
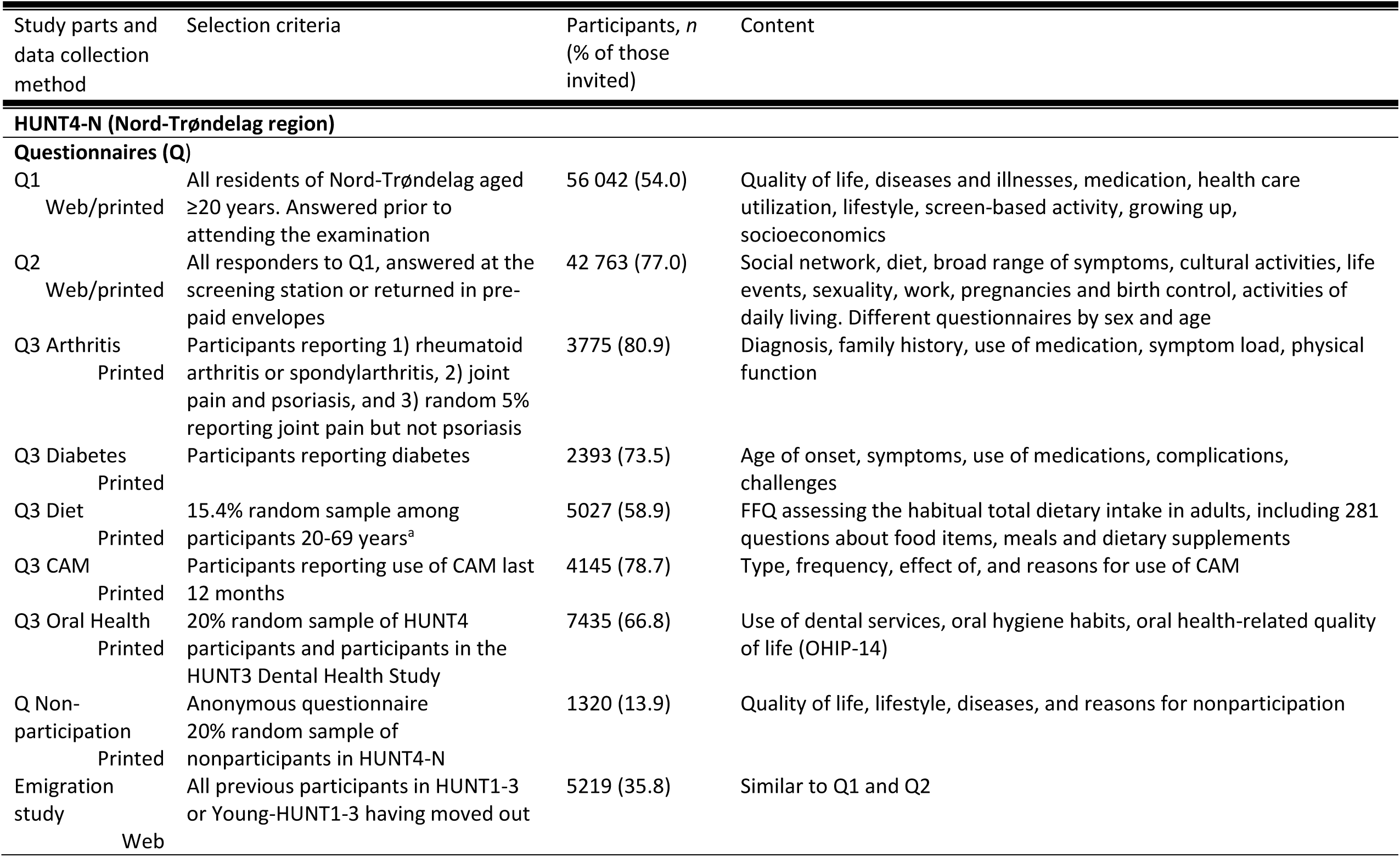
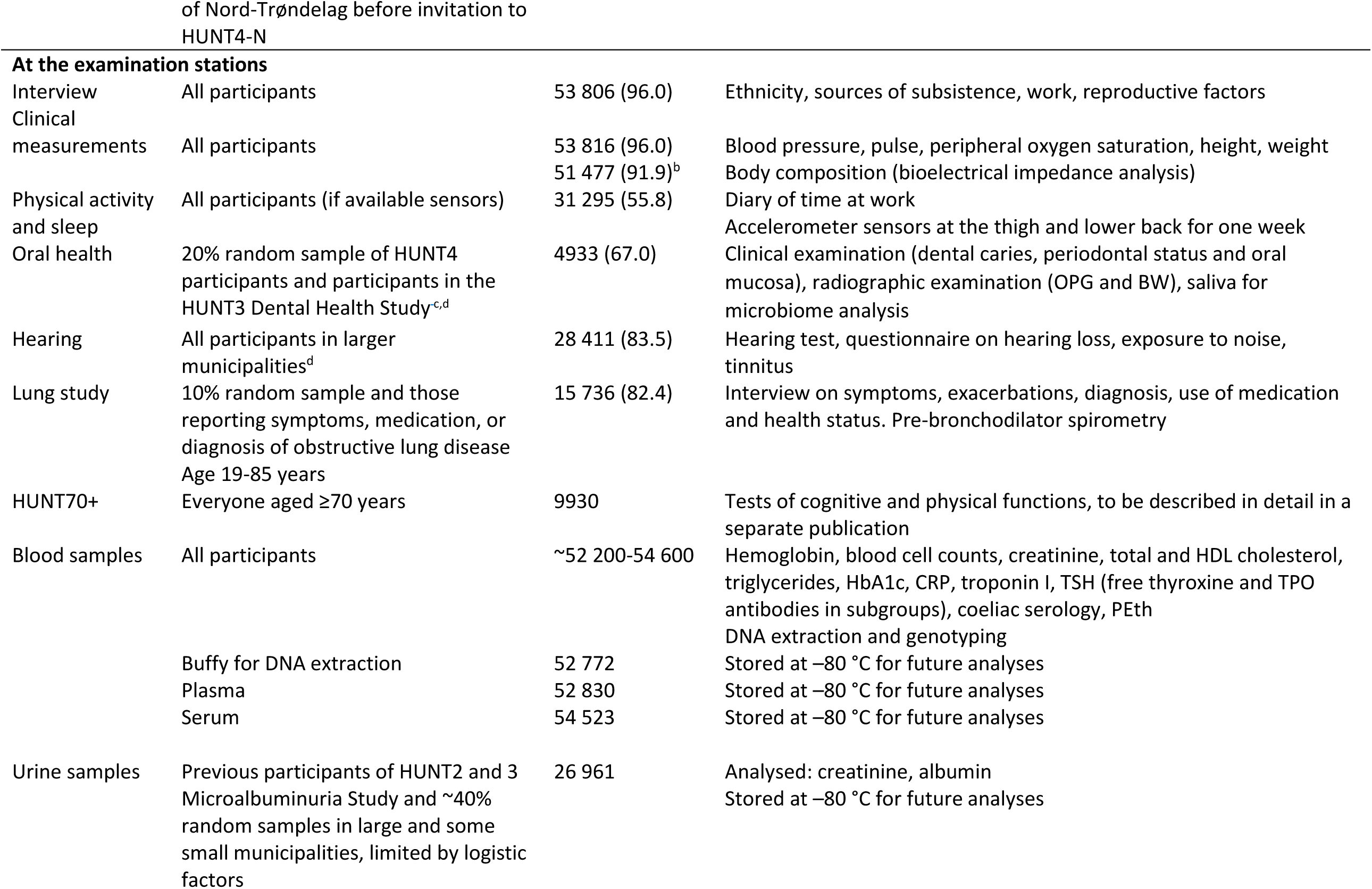
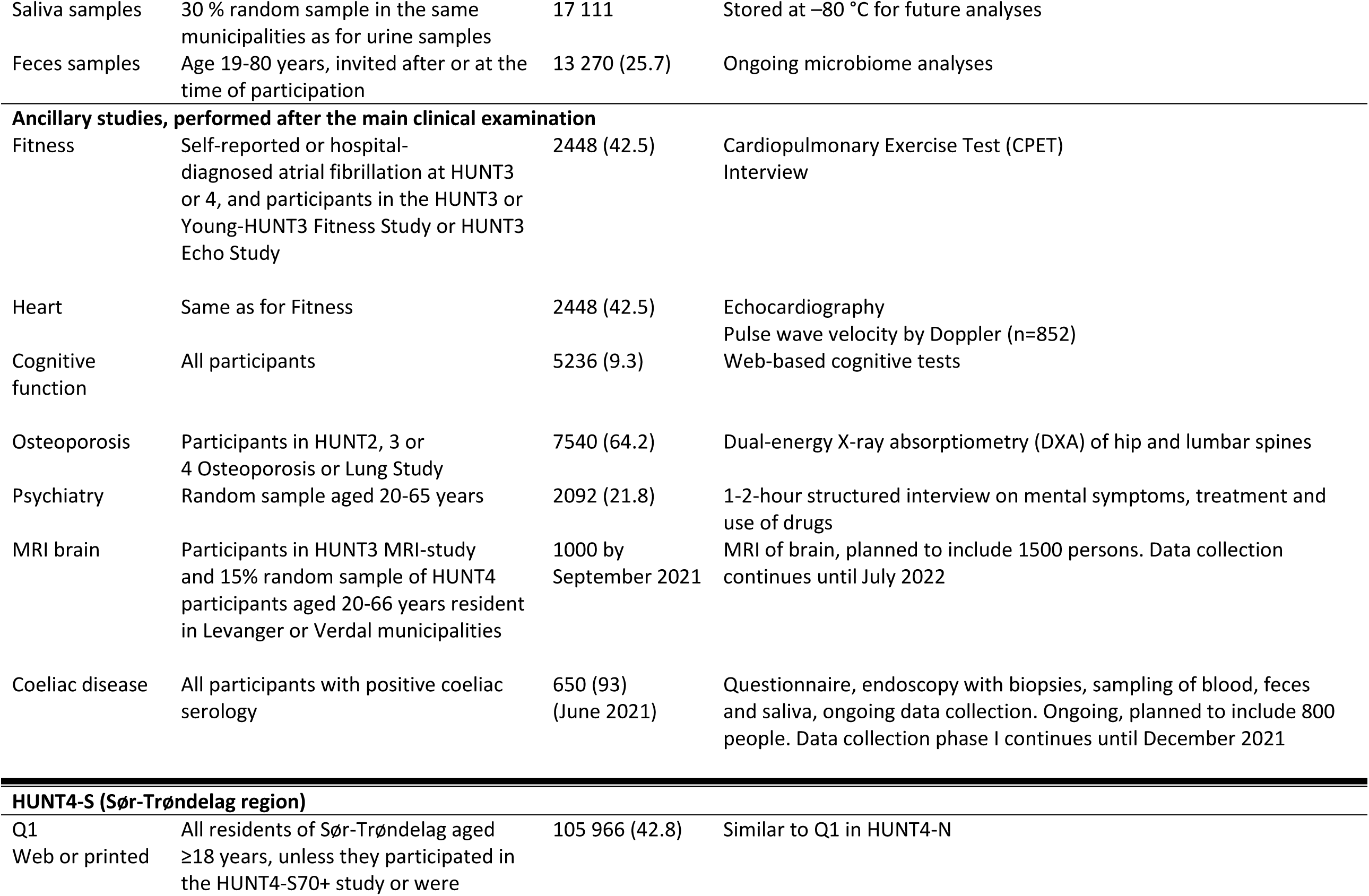
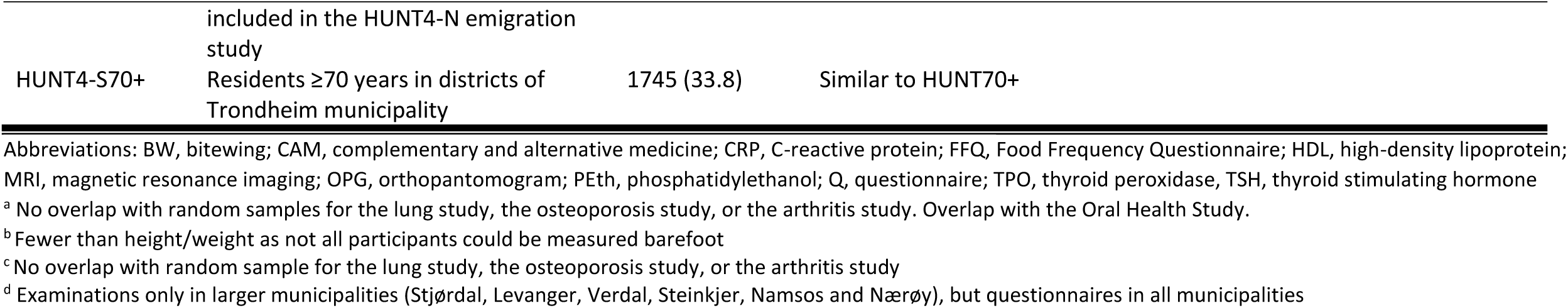
Content of the HUNT4-N and HUNT4-S surveys.

### What has been measured?

Similar to previous HUNT surveys, HUNT4-N consisted of questionnaires, a short interview, clinical examination, and biological sampling, and was conducted by trained health professionals at examination stations in each of 23 municipalities in Nord-Trøndelag. HUNT4-S was mainly a web-based questionnaire survey (except for the examination of 1745 residents ≥70 years of age in Trondheim), but printed questionnaires were sent on request and to older adults. The HUNT4 questionnaires covered a broad range of topics including socio-economic conditions, health-related behaviors, symptoms, conditions and diseases, as described in Table 3. Selected HUNT4-N participants were also asked to complete questionnaires related to specific health topics, and a 15% random sample aged 20-69 years was invited to complete a web-based food frequency questionnaire.^5^

An overview of HUNT4 measurements, performed as part of the main HUNT4 examination or as ancillary data collections, is provided in Table 3. New aspects of the clinical examination in HUNT4-N compared to previous HUNT surveys included detailed body composition analysis using bioelectrical impedance (InBody 770, Cerritos, CA, USA), a one-week accelerometer recording (AX3, Axivity Ltd., Newcastle, UK), and physical and cognitive testing (Short Physical Performance Battery (SPPB), grip strength and Montreal Cognitive Assessment, MoCA; participants ≥70 years of age). Blood pressure, pulse and peripheral capillary oxygen saturation were recorded three times at one-minute intervals using Dinamap CARESCAPE V100 (GE Healthcare, Chicago, Illinois, USA). Oral health clinical and radiographic examinations (Planmeca ProOne (orthopantomogram) and Planmeca Intra / Prostyle Intra with ProSensor HD (bitewing), Helsinki, Finland), hearing test (air-conduction pure-tone audiometry at 0.25-8 kHz according to ISO 8253-1 using Interacoustics audiometers type AD629 with TDH-39P supra-aural audiometric earphones) and spirometry (Jaeger Masterscope spirometers, JLAB version latest upgrade 2016, CareFusion, Würzburg, Germany) were performed in selected samples, and other tests have subsequently been performed in subgroups. All participants provided blood samples drawn in a non-fasting state between 9 am and 8 pm and time since last meal was recorded. Biological sampling at the field stations included blood, urine and saliva, while feces kits were returned in pre-paid envelopes. Biologic material was handled at the field stations according to appropriate standards and transported to the biobank every evening in a cold chain. For all participants, two blood tubes were delivered at the laboratory of Levanger Hospital, Nord-Trøndelag Hospital Trust the next day for immediate analyses or transport to other laboratories. New analyses in blood included hemoglobin (Hb) and blood cell counts, HbA1c and phosphatidylethanol (PEth, an indicator of alcohol intake). Three blood tubes were aliquoted and stored in automated freezers in HUNT Biobank.

Since the publication of the original cohort profile in 2013,^3^ new analyses of biological material stored in HUNT Biobank include the genotyping of 70 517 participants of HUNT2 or HUNT3, genotyping of 18 722 additional participants of HUNT4-N (details on genotyping and imputation will be described in a separate paper), and SomaLogic’s SomaScan proteomics analyses^6^ and measurements of vitamin D^7^ and troponin I^8^ in subsamples.

### What has it found? Key findings and publications

In Table 1, we present descriptive characteristics of the HUNT4-N and HUNT4-S participants, including prevalence estimates of a range of chronic conditions and diseases. The longitudinal nature of HUNT enables analyses of long-term population changes in health-related factors. For example, comparison of HUNT4-N with previous HUNT surveys shows how the prevalence of smoking, high blood pressure and atherogenic lipid levels has declined from the 1980s until now, whereas the prevalence of obesity and diabetes has increased (Figure 2, Supplementary Table S4). Although the prevalence of diabetes has increased, HbA1c measurements in HUNT4 indicate that the current prevalence of undiagnosed diabetes is low.^9^ The prevalence of tension-type headache has increased, whereas migraine and medication overuse headache have become less common,^10^ as has hearing impairment.^11^ Information from HUNT4 has further enabled prevalence estimates of dementia, mild cognitive impairment,^12^ periodontitis^13^ and depression and anxiety symptoms,^14^ and estimation of the longitudinal decline in VO_2peak_.^15^ Analyses of HUNT4 have further shown that shift work is associated with higher levels of C-reactive protein and chronic musculoskeletal pain,^16^ and that cognitive impairment is associated with lower physical performance.^17^

**Figure 2.**
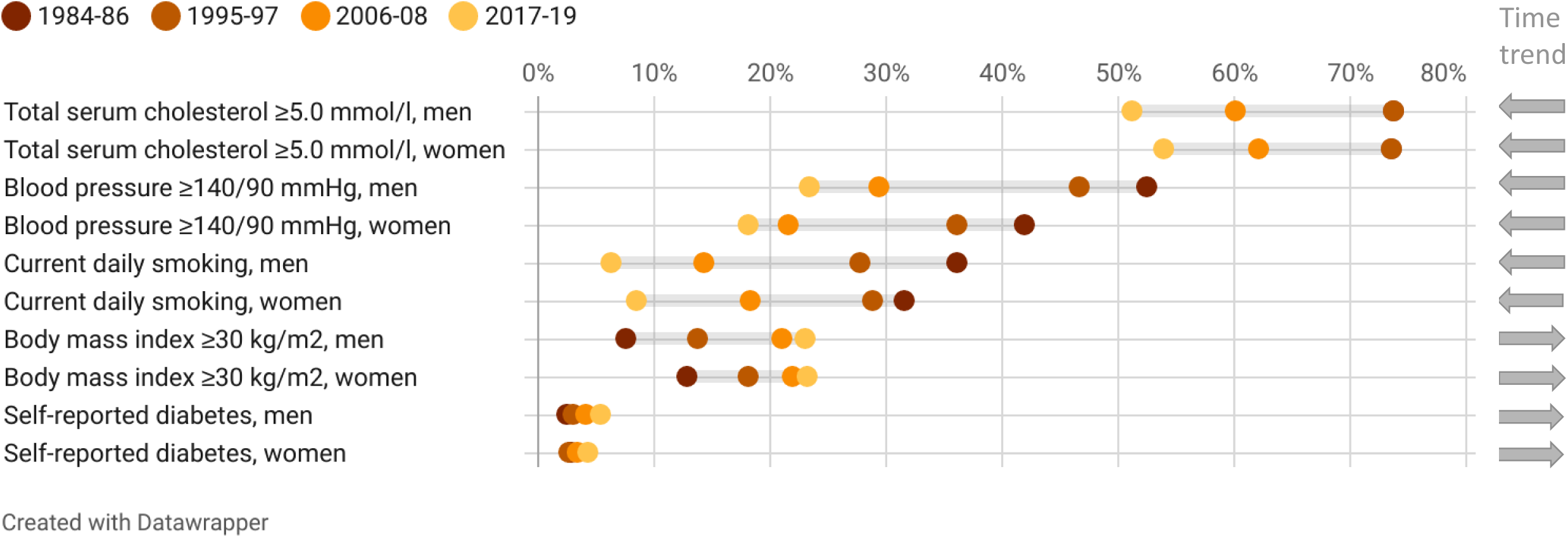
Prevalence of cardiovascular risk factors in HUNT1 (1984-86), HUNT2 (1995-97), HUNT3 (2006-08) and HUNT4-N (2017-19), by sex, age-standardized (direct method using 10-year age categories) to the Norwegian population on January 1, 2020. Point estimates with 95% confidence intervals are provided in Supplementary Table S4.

The new focus on genetic and molecular epidemiology has resulted in e.g. genome-wide association studies (GWAS) identifying new genetic variants associated with atrial fibrillation,^18^ serum lipids,^19^ thyroid stimulating hormone (TSH)^20^ and bone mineral density.^21^ Mendelian randomization studies have examined the causal associations between serum lipids and kidney function^22^ and provided support to the causal role of smoking heaviness on mortality,^23^ of higher body mass on psoriasis^24^ and bloodstream infections,^25^ and of PCSK9 on serum cholesterol and coronary heart disease.^26^ Anthropometric measures spanning six decades have demonstrated how body mass index (BMI) increases more strongly in genetically predisposed individuals during transition to a more obesogenic environment.^27, 28^ The predictive value of circulating proteins on secondary cardiovascular events has been examined using proteomics analyses.^6^ The large number of relatives participating in HUNT has enabled within-family studies. Parent-offspring analyses have suggested that maternal intrauterine environment, as proxied by maternal SNPs that influence offspring birthweight, is unlikely to be a major determinant of adverse cardiometabolic outcomes in population-based samples of offspring.^29^ Within-family Mendelian randomization analyses have enabled correction for familial biases in e.g. studies of the effects of height and body mass on educational attainment.^30^ Supplementary Table S5 provides examples of studies showing how HUNT data have been used across a range of health topics since the original cohort profile was published in 2013.

### What are the main strengths and weaknesses?

HUNT is suitable for longitudinal studies due to the long-term follow-up with repeated measurements since the 1980s, low migration, and available valid information on migration and deaths from the National Population Register. The HUNT surveys cover a broad range of health-related topics, and many questionnaire items have been kept unchanged across the surveys to enable longitudinal comparisons. Quality-controlled HUNT data are stored at the HUNT Databank and biological material is stored in the state-of-the-art HUNT Biobank at the HUNT Research Centre, Levanger. The HUNT participants have consented to linkage to the many high-quality health and administrative registries in Norway, and to information from medical records. Such linkages can be reliably made using the national identification numbers allocated to all Norwegian residents and means that prospectively recorded information on health outcomes can be obtained also for participants who do not attend subsequent HUNT surveys. The ethnically homogenous population limits the generalizability to people of non-European ancestry.

As the HUNT Study has invited the entire population in the area over decades, the study population includes many family members, both siblings and across generations, and is therefore suitable for within-family studies. A particular asset compared with many other studies is available offspring information in adult age.^31-33^ The study population has cryptic relatedness that may have to be considered in the analytical approach of e.g. GWAS but is quite outbred with a mean inbreeding coefficient^34^ of 0.0024 (calculated using KING software^35^).

The relatively high participation indicates a generally lower concern for selection bias. Nevertheless, attrition from HUNT3 to HUNT4 was somewhat higher among participants with chronic diseases or other indices of ill health, and lifestyle differed among nonparticipants and migrants compared with participants of HUNT4. To inform about selective participation by ill health among all HUNT4 invitees, we also examined primary health care diagnostic codes across a range of diseases, as well as primary health care utilization, for HUNT4 invitees and participants recorded during the calendar year 2017 (for HUNT4-N) or 2019 (for HUNT4-S). The proportion being assigned each diagnostic code generally did not differ substantially between participants and invitees, but results vary between diagnoses as detailed in Supplementary Table S6. For example, a dementia diagnosis was less often recorded among participants than among invitees. General practice visits were more frequent among participants than nonparticipants at ages >80 years. In contrast, home nursing and nursing home residency were more common among nonparticipants (Supplementary Figure S1).

To evaluate the quality of the self-report of conditions in HUNT4, we compared the self-reported information to diagnostic codes recorded in the local or regional hospitals and on general practitioners’ reimbursement forms. Compared to these diagnoses as a reference standard, the sensitivity, specificity and predictive values of the self-reported information varied across diagnoses in both HUNT4-N (Supplementary Table S7) and HUNT4-S (Supplementary Table S8). Of note, the diagnostic codes may be inaccurate and do not constitute a definite reference standard. For example, in lack of a more suitable diagnostic code, a disease diagnosis may likely be reported on the reimbursement form by a general practitioner if a patient comes for testing for that disease, even if the disease was not confirmed. In-depth validity studies have been conducted for self-reported headache^36^ and insomnia^37^ in HUNT4. Validity studies of self-report of e.g. diabetes,^38^ psoriasis^39^ and atrial fibrillation^40^ have been performed after previous HUNT surveys.

### Can I get hold of the data? Where can I find out more?

Researchers affiliated to a Norwegian research institution can apply for HUNT data access from HUNT Research Centre (www.ntnu.edu/hunt) if they have obtained project approval from the Regional Committee for Medical and Health Research Ethics (REC). Researchers not affiliated to a Norwegian research institution should collaborate with and apply through a Norwegian principal investigator. Information on the application and conditions for data access is available at www.ntnu.edu/hunt/data. The HUNT Databank website provides a detailed overview of the available variables in HUNT (www.ntnu.edu/hunt/databank). Certain data from ancillary HUNT projects may be subject to a time-limited exclusivity provided to the researchers who have financed and conducted the data collection. Biologic material is available for analyses, information on procedures is found at the HUNT Biobank website (https://www.ntnu.edu/hunt/hunt-biobank). Data from the health registries are not kept by HUNT; instead, linkages between HUNT and registry data have to be made for each research project and require that the principal investigator has obtained project-specific approval for such linkage by REC and each registry owner.

## Data Availability

HUNT data availability is described in the "Can I get hold of the data? Where can I find out more?" section of the manuscript.

## Acknowledgements

The Trøndelag Health Study (HUNT) is a collaboration between HUNT Research Centre (Faculty of Medicine and Health Sciences, NTNU, Norwegian University of Science and Technology), Trøndelag County Council, Central Norway Regional Health Authority, and the Norwegian Institute of Public Health. We thank the population of Trøndelag for their willingness to contribute with important data and biologic material, and politicians and administrations in all municipalities for positive attitude and support of logistics. We thank the administrative staff at HUNT Research Centre for comprehensive contribution to the planning, performance, collection and storage of data and biologic material. For this Cohort Profile Update, Nord-Trøndelag Hospital Trust provided diagnostic codes from Nord-Trøndelag Hospital Trust and St. Olavs hospital, and the Norwegian Directorate of Health provided data from the KUHR database and the Norwegian Registry for Primary Health Care. The interpretation and reporting of these data are the sole responsibility of the authors, and no endorsement by the Norwegian Registry for Primary Health Care is intended nor should be inferred. The work presented in this Cohort Profile Update was approved by the Mid-Norway Regional Committee for Medical and Health Research Ethics (REK midt 67445).

