## Supplementary Table S1 for "Cohort Profile Update: The HUNT Study, Norway"

**Supplementary Table S1.** Comparisons of demographics, dispensed medication, cancer rates, and causes of death for 2018 between the populations of Trøndelag[a] and Norway.

| | Trøndelag | Norway |
|---|---|---|
| Number of inhabitants, January 1, 2018 | 468 702 | 5 295 619 |
| **Demographics, socioeconomics and lifestyle data[b]** (in percent unless otherwise stated) | | |
| Proportion of the population aged 0-17 years | 20.8 | 21.1 |
| Predicted proportion of the population aged ≥80 years in 2025 | 4.3 | 4.2 |
| Education ≥12 years among people aged 30-39 years | 82 | 80 |
| Low income (households)[c] | 6.5 | 7.4 |
| Social benefits for daily living among people aged 20-66 years | 16.0 | 15.6 |
| Air quality, fine dust (µg/m$^3$) | 4.1 | 5.4 |
| Reported violence to police (per 1000) | 5.3 | 7.0 |
| Hip fractures, aged ≥75 years (per 1000) | 17 | 17 |
| Daily smokers among people aged 16-44 years | 5.7 | 6.9 |
| Daily smokers among people aged 45-74 years | 14.0 | 14.0 |
| Life expectancy among men (years) | 80.8 | 80.4 |
| Life expectancy among women (years) | 84.3 | 84.0 |
| **Dispensed prescription medication (Anatomical Therapeutic Chemical [ATC] codes) per 1000 persons[d]** | | |
| A02BA Proton pump inhibitors | 88.6 | 96.0 |
| A10A Insulin, insulin analoges | 13.1 | 12.8 |
| A10B Blood glucose lowering drugs, excluding insulin | 29.3 | 30.3 |
| C Cardiovascular system | 205.5 | 213.4 |
| C07 Beta-blockers | 61.9 | 71.4 |
| C10 Lipid lowering drugs | 99.6 | 109.6 |
| B01 Antithrombotic drugs | 105.9 | 104.8 |
| R03 Medications for obstructive airway disease | 78.8 | 86.9 |
| **Cancer, age-standardized rates per 100,000 person-years[e]** | | |
| All types, women | 523.7 | 548.8 |
| All types, men | 657.3 | 700.0 |
| Breast, women | 123.7 | 128.6 |
| Central nervous system, women | 19.2 | 16.1 |
| Central nervous system, men | 17.0 | 16.5 |
| Gastrointestinal tract, women | 100.3 | 105.8 |
| Gastrointestinal tract, men | 144.8 | 145.8 |
| Airways, women | 55.6 | 57.7 |

| | | |
|---|---|---|
| Airways, men | 57.8 | 68.9 |
| Urinary tract, women | 23.8 | 26.4 |
| Urinary tract, men | 66.2 | 71.1 |
| Genitalia, women | 60.1 | 61.9 |
| Genitalia, men | 176.8 | 194.4 |

| **Causes of death (% of deaths in 2018)[f]** | | |
|---|---|---|
| Total number | 3546 | 40 786 |
| Infectious diseases | 2.1 | 2.3 |
| Tumors | 27.4 | 27.3 |
| Mental diseases | 7.3 | 7.8 |
| Nervous system diseases | 5.0 | 5.5 |
| Cardiovascular diseases | 25.4 | 24.7 |
| Respiratory diseases | 9.9 | 11.0 |
| Gastrointestinal diseases | 2.7 | 3.0 |

[a] The counties of Nord-Trøndelag and Sør-Trøndelag were merged on January 1, 2018. Thereafter national statistics are given for the merged Trøndelag county. The main demographic difference between the two previous counties is the location of a large city in Sør-Trøndelag (Trondheim with about 202 000 inhabitants (2019)), compared to smaller cities with 14-22 000 inhabitants in Nord-Trøndelag. According to the Health Profile for 2017, Nord-Trøndelag compared to Sør-Trøndelag, will have higher predicted proportion aged ≥80 years in 2025 (5.7 versus 4.6%), has a lower proportion with education ≥12 years (81 versus 86% among people aged 30-39 years), and a higher proportion of smokers among people aged 16-44 years (9.6 versus 6.2%), but otherwise, the populations had similar characteristics.

[b] Norwegian Institute of Public Health, Public Health Profile 2020, page 4. https://www.fhi.no/hn/folkehelse/folkehelseprofil/

[c] All ages, persons living in households with income <60% of national median (about €30,000) and gross financial capital <€10,000

[d] The Norwegian Prescription Database (NorPD), https://www.norpd.no

[e] The Cancer Registry of Norway, https://sb.kreftregisteret.no/insidens/?lang=en

[f] Norway Cause of Death Registry, http://ghdx.healthdata.org/series/norway-cause-death-registry
