## Supplementary Table S2 for "Cohort Profile Update: The HUNT Study, Norway"

**Supplementary Table S2.** Relative probability of participation in HUNT4-N according to characteristics at participation in HUNT3.[a]

| Characteristics at participation in HUNT3 (2006-08) | Women | | Men | |
|---|---|---|---|---|
| | HUNT4-N participants, *n* (%)[b] | Relative probability[c] (95% CI) | HUNT4-N participants, *n* (%)[b] | Relative probability[c] (95% CI) |
| Age (years) | | | | |
| 19-29 | 1442 (54.5) | 1.00 (reference) | 920 (49.3) | 1.00 (reference) |
| 30-39 | 3029 (75.6) | 1.39 (1.33-1.44) | 1923 (67.4) | 1.37 (1.30-1.44) |
| 40-49 | 4337 (79.6) | 1.46 (1.41-1.52) | 3390 (74.3) | 1.51 (1.43-1.58) |
| 50-59 | 4802 (80.0) | 1.47 (1.41-1.52) | 4211 (77.7) | 1.58 (1.50-1.65) |
| 60-69 | 3670 (71.6) | 1.31 (1.26-1.37) | 3193 (68.3) | 1.39 (1.32-1.46) |
| 70-79 | 1459 (47.3) | 0.87 (0.82-0.91) | 1106 (41.5) | 0.84 (0.79-0.90) |
| ≥80 | 217 (15.1) | 0.28 (0.24-0.31) | 120 (12.0) | 0.24 (0.20-0.29) |
| Marital status | | | | |
| Married or registered partner | 11 722 (75.6) | 1.00 (reference) | 9778 (68.7) | 1.00 (reference) |
| Not married or registered partner | 3801 (64.4) | 0.90 (0.88-0.92) | 3489 (59.7) | 0.86 (0.84-0.89) |
| Divorced, separated or widow(er) | 3422 (54.2) | 0.87 (0.85-0.89) | 1583 (54.0) | 0.82 (0.80-0.85) |
| Constituency type | | | | |
| Urban | 6758 (67.6) | 1.00 (reference) | 5043 (64.5) | 1.00 (reference) |
| Rural | 11 599 (70.1) | 1.03 (1.01-1.04) | 9438 (65.9) | 1.02 (1.00-1.04) |
| Body mass index (kg/m$^2$) | | | | |
| <25.0 | 7551 (69.8) | 1.00 (reference) | 3553 (60.5) | 1.00 (reference) |
| 25.0-29.9 | 7267 (70.1) | 1.00 (0.99-1.02) | 8127 (68.0) | 1.05 (1.03-1.07) |
| ≥30.0 | 4072 (64.0) | 0.92 (0.90-0.94) | 3127 (62.1) | 0.95 (0.92-0.97) |
| Good or very good self-rated health | | | | |
| Yes | 13 943 (72.8) | 1.00 (reference) | 11 726 (68.1) | 1.00 (reference) |
| No | 4421 (58.1) | 0.86 (0.84-0.88) | 2807 (53.4) | 0.85 (0.83-0.88) |
| Daily smoking | | | | |
| Never | 9551 (69.3) | 1.00 (reference) | 7627 (69.8) | 1.00 (reference) |
| Former | 5763 (72.4) | 0.99 (0.97-1.00) | 5071 (62.2) | 0.94 (0.92-0.96) |
| Current | 3185 (61.5) | 0.81 (0.79-0.83) | 1829 (54.1) | 0.77 (0.74-0.79) |
| History of current or previous disease (self-reported) | | | | |
| Coronary heart disease (myocardial infarction or angina pectoris) | 18 544 (69.4) | | | |
| No | 18 544 (69.4) | 1.00 (reference) | 13 965 (66.4) | 1.00 (reference) |

| | | | | |
|---|---|---|---|---|
| Yes | 410 (39.3) | 0.83 (0.78-0.89) | 896 (44.9) | 0.85 (0.81-0.89) |
| Stroke | | | | |
| No | 18 657 (68.9) | 1.00 (reference) | 14 564 (65.3) | 1.00 (reference) |
| Yes | 297 (44.9) | 0.83 (0.77-0.90) | 297 (40.7) | 0.78 (0.72-0.84) |
| Asthma | | | | |
| No | 16 762 (68.8) | 1.00 (reference) | 13 287 (64.9) | 1.00 (reference) |
| Yes | 2192 (64.9) | 0.95 (0.93-0.97) | 1574 (61.2) | 0.96 (0.93-0.99) |
| COPD or emphysema | | | | |
| No | 18 478 (68.9) | 1.00 (reference) | 14 508 (65.3) | 1.00 (reference) |
| Yes | 473 (50.8) | 0.78 (0.74-0.83) | 352 (43.3) | 0.76 (0.70-0.82) |
| Diabetes | | | | |
| No | 18 387 (69.0) | 1.00 (reference) | 14 273 (65.3) | 1.00 (reference) |
| Yes | 566 (51.6) | 0.87 (0.83-0.92) | 586 (50.2) | 0.83 (0.79-0.88) |
| Cancer | | | | |
| No | 18 046 (69.1) | 1.00 (reference) | 14 343 (65.6) | 1.00 (reference) |
| Yes | 907 (55.1) | 0.90 (0.86-0.93) | 518 (44.8) | 0.83 (0.78-0.88) |
| Sought health care for mental health problems | | | | |
| No | 14 906 (69.6) | 1.00 (reference) | 12 850 (65.4) | 1.00 (reference) |
| Yes | 3446 (65.3) | 0.92 (0.90-0.93) | 1617 (61.5) | 0.91 (0.89-0.94) |
| Visit to a general practitioner during the last 12 months | | | | |
| No | 3163 (70.6) | 1.00 (reference) | 4031 (68.5) | 1.00 (reference) |
| Yes | 15 792 (67.9) | 1.00 (0.98-1.02) | 10 830 (63.1) | 0.99 (0.97-1.01) |
| Admitted to hospital during the last 12 months | | | | |
| No | 16 828 (69.2) | 1.00 (reference) | 13 500 (65.8) | 1.00 (reference) |
| Yes | 2127 (61.8) | 0.94 (0.92-0.96) | 1362 (54.2) | 0.89 (0.86-0.92) |

[a] In total, 18 956 (68.3%) of 27 754 female HUNT3 participants and 14 863 (64.5%) of 23 046 male HUNT3 participants participated in HUNT4; the remaining HUNT3 participants either died or moved out of Nord-Trøndelag between the HUNT3 and HUNT4 surveys or were invited but did not participate in HUNT4.

[b] Percentage among the total number of HUNT3 participants in the category

[c] Relative probabilities of participation, adjusted for age (in 10-year categories) at participation in HUNT3
