## Supplementary Table S3 for "Cohort Profile Update: The HUNT Study, Norway"

**Supplementary Table S3.** Characteristics of participants and nonparticipants of HUNT4-N, and previous HUNT participants having emigrated from Nord-Trøndelag. Percentages based on self-reported information unless otherwise noted. Age-standardized to the 10-year age group distribution in HUNT4-N.

| Characteristics | HUNT4-N participants | | Invited to HUNT4-N, but did not participate | | Emigrated from Nord-Trøndelag | |
|---|---|---|---|---|---|---|
| | Women | Men | Women | Men | Women | Men |
| Number of participants (% of invitees) | 30 574 (54.6) | 25 468 (45.4) | 649 (49.0) | 671 (51.0) | 3061 (55.8) | 2421 (44.2) |
| Age (years), mean | 53.9 | 54.8 | 53.4 | 54.4 | 54.2 | 55.1 |
| Body mass index ($kg/m^2$), mean[a] | 26.9 | 27.6 | 27.4 | 27.5 | 25.5 | 26.6 |
| Physical exercise $\geq$2-3 times/week | 67.6 | 59.4 | 54.0 | 50.2 | 75.9 | 73.0 |
| Current daily smoking | 9.6 | 7.0 | 13.5 | 11.4 | 6.6 | 4.7 |
| Never smoking | 56.1 | 55.9 | 54.4 | 47.7 | 46.4 | 47.0 |
| Alcohol intake $\geq$2-3 times/week | 15.1 | 25.0 | 11.0 | 21.0 | 25.4 | 39.4 |
| Good or very good self-rated health | 73.6 | 78.6 | 67.5 | 73.4 | 77.7 | 82.7 |
| Current use of antihypertensive medication | 21.4 | 25.6 | 28.1 | 30.0 | 17.8 | 28.4 |
| History of | | | | | | |
| Myocardial infarction /angina | 3.5 | 8.6 | 4.2 | 9.8 | 2.5 | 4.3 |
| Stroke | 2.9 | 4.1 | 3.4 | 4.6 | 2.3 | 4.3 |
| Asthma | 12.5 | 11.6 | 14.3 | 10.0 | 12.1 | 10.7 |
| COPD | 2.6 | 3.3 | 4.8 | 4.1 | 1.8 | 3.6 |
| Diabetes | 5.2 | 7.1 | 7.5 | 11.4 | 4.1 | 8.1 |
| Hypo-or hyperthyroidism | 12.1 | 3.3 | 14.8 | 5.3 | 12.8 | 3.9 |
| Cancer | 7.9 | 8.0 | 9.8 | 8.1 | 9.3 | 10.5 |
| Sought health care for mental health problems | 21.6 | 12.2 | 23.6 | 12.2 | 25.0 | 12.4 |

[a]Height and weight measured in HUNT4-N, otherwise self-reported
