## Supplementary Table S4 for "Cohort Profile Update: The HUNT Study, Norway"

**Supplementary Table S4.** Prevalence with 95% confidence interval (CI) of cardiovascular risk factors in HUNT1 (1984-86), HUNT2 (1995-97), HUNT3 (2006-08) and HUNT4-N (2017-19), by sex, age-standardized (direct method using 10-year age categories) to the Norwegian population on January 1, 2020.

| | 1984-86 | | 1995-97 | | 2006-08 | | 2017-19 | |
|---|---|---|---|---|---|---|---|---|
| | Percent | 95% CI | Percent | 95% CI | Percent | 95% CI | Percent | 95% CI |
| Total serum cholesterol ≥5.0 mmol/l, men | | | 73.8 | 73.3-74.3 | 60.3 | 59.6-61.0 | 51.4 | 50.7-52.0 |
| Total serum cholesterol ≥5.0 mmol/l, women | | | 73.7 | 73.2-74.1 | 62.3 | 61.7-62.9 | 54.0 | 53.4-54.5 |
| Blood pressure ≥140/90 mmHg, men | 52.5 | 52.0-53.0 | 46.8 | 46.3-47.4 | 29.5 | 28.9-30.1 | 23.4 | 23.0-23.9 |
| Blood pressure ≥140/90 mmHg, women | 42.0 | 41.6-42.4 | 36.3 | 35.9-36.7 | 21.7 | 21.3-22.1 | 18.3 | 18.0-18.7 |
| Current daily smoking, men | 36.3 | 35.8-36.9 | 28.0 | 27.4-28.5 | 14.4 | 13.9-14.9 | 6.5 | 6.2-6.8 |
| Current daily smoking, women | 31.6 | 31.1-32.1 | 29.0 | 28.6-29.5 | 18.5 | 18.0-19.0 | 8.6 | 8.3-8.9 |
| Body mass index ≥30 $kg/m^2$, men | 7.7 | 7.4-7.9 | 13.9 | 13.5-14.3 | 21.1 | 20.5-21.7 | 23.2 | 22.7-23.8 |
| Body mass index ≥30 $kg/m^2$, women | 13.0 | 12.7-13.4 | 18.2 | 17.8-18.6 | 22.2 | 21.7-22.7 | 23.3 | 22.8-23.9 |
| Self-reported diabetes, men | 2.7 | 2.6-2.9 | 3.2 | 3.0-3.4 | 4.2 | 4.0-4.5 | 5.5 | 5.2-5.7 |
| Self-reported diabetes, women | 3.0 | 2.8-3.2 | 2.9 | 2.7-3.1 | 3.5 | 3.3-3.7 | 4.4 | 4.2-4.6 |
