## Supplementary Table S5 for "Cohort Profile Update: The HUNT Study, Norway"

**Supplementary Table S5.** Examples on how HUNT data have been used across a range of health topics since the original cohort profile was published in 2013.

| Topic | Finding/contribution | PubMed IDentifier (PMID) |
| --- | --- | --- |
| Airways | Influenced change in the international COPD classification (ABCD).<br>Validated Global Lung Initiative reference values, now included in Norway. Diagnostic criteria for COPD changed accordingly.<br>Updated Norwegian prevalence data and trends of COPD<br>Certain comorbidity clusters, as identified using machine learning software, are associated with increased mortality and risk of severe exacerbations in COPD (under review).<br>A century of increasing lung function and its implications for the diagnosis of lung disease: Results from 243,465 European adults across ten population-based studies<br>Contribution to international consortia on lung function reference values and lung cancer risk factors | 23611880<br>27824594<br>30180485<br>29724393<br>22743675<br>29617726 |
| Biometrics | A century of trends in adult human height | 27458798 |
| Bone health | Osteoporosis is associated with increased mortality in individuals with COPD.<br>Underuse of anti-osteoporotic treatment in persons with high fracture risk<br>The FRAX risk score without bone mineral density predicts hip fractures reasonably well.<br>HUNT- data included in ongoing update of FRAX (FRAX2). | 31709837<br>29774403<br>28668994 |
| Cardiovascular system (CVD): Blood pressure | Worldwide trends in blood pressure from 1975 to 2015<br>Contributions of mean and shape of blood pressure distribution to worldwide trends and variations in raised blood pressure | 27863813<br>29579276 |
| CVD:<br>Atrial Fibrillation | Estimated cardiorespiratory fitness inversely associated with AF<br>Higher physical activity levels and fitness associated with lower risk of CVD and all-cause mortality<br>Physical activity modifies AF risk in obese | 31246716<br>32047884<br>29939081 |
| CVD: Stroke | Albuminuria is a risk factor for ischemic stroke.<br>The risk of stroke depends on the metabolic consequences of obesity. | 32359353<br>34281375 |
| Cardiorespiratory fitness | Validated nonexercise model of fitness for prediction of mortality and morbidity.<br>Provided data on the link between fitness and coronary disease. | 24576863<br>30496487 |

| | | |
|---|---|---|
| | New data linking fitness to cardiac structure and function.<br>Creation and validation of Physical Activity Intelligence index (PAI) to motivate people to become and mainstay physical healthy.<br>Physical activity has protective effect on depression after myocardial infarction. | 31986991<br>32971113<br>27866655<br>27984009<br><br>26302141 |
| Cardiovascular: Echocardiography | Largest normal reference ranges for advanced echocardiography.<br><br>Cardiac function of healthy individuals is impaired by risk factors for cardiac disease.<br>Established how cardiac geometry influence measures of cardiac function.<br>Evaluated important factors for test-retest variation of echocardiographic measurements<br>Creation of optimally matched control groups for evaluation of cardiotoxicity in cancer patients.<br><br>Creation of optimally matched control groups for evaluation of cardiac dysfunction diabetic patients.<br>Depression symptoms influence cardiac function. | 19946115<br>20581050<br>31544286<br>21247733<br><br>32154940<br>32978265<br>33960012<br>19959533<br>27038515<br>26897666<br>26169610<br>26948543<br>25350248<br>26925243 |
| Cardiovascular: General | Cardiovascular outcome is impaired by symptoms of depression and anxiety.<br>Diabetes influence mortality different in men and women.<br>Reduced cardiovascular mortality in diabetic patients between HUNT1 and HUNT3.<br>Established novel genes important for cardiac diseases.<br>Pregnancy complications predicts cardiovascular diseases.<br>Cardiac biomarkers predict cardiovascular prognosis. | 24057077<br>25044493<br>17947212<br><br>18595902<br><br>24633158<br>29290336<br>30596987<br>31188397<br>27815376<br>26294790<br>30996050 |
| Chronic pain and sleep | Insomnia symptoms are associated with increased risk of chronic pain and pain-related disability. A physically active lifestyle may to some extent compensate the adverse effect of insomnia symptoms on risk of chronic pain. | 24293504<br>28744933<br>29699540<br>31801790 |
| Dementia | Current and future prevalence estimates of mild cognitive impairment, dementia, and its subtypes<br>Physical performance across the cognitive spectrum and between dementia subtypes<br>Temporal changes in cardiorespiratory fitness and risk of dementia incidence and mortality<br>C-reactive protein, blood pressure and chronic kidney disease as risk factors for incident dementia | 33427745<br><br>33798998<br><br>31677775<br><br>29387136<br>28569205<br>31299931 |

| | | |
|---|---|---|
| Diabetes | Worldwide trends in diabetes since 1980<br>Low C-peptide and high glutamic acid decarboxylase autoantibody levels predict progression to insulin dependence in LADA<br>High physical activity level is associated with reduced risk of LADA in individuals without high-risk HLA genetic susceptibility<br>Overweight interacts with HLA high-risk genotypes but also with genes associated with type 2 diabetes in the promotion of LADA<br>The validity of FINDRISC and the risk of diabetes among people with FINDRISC ≥15 is lower than assumed in national guidelines<br>A subset of individuals fulfilling diagnostic criteria for type 2 diabetes display transient signs of autoimmunity preceding diagnosis | 27061677<br>34318969<br>32835373<br>31125083<br>31803483<br>30327361 |
| Eating disorders | Maternal eating disorders are associated with adverse obstetric outcomes. | 30189108 |
| Family studies | Parental chronic musculoskeletal pain is associated with increased risk of chronic musculoskeletal pain in the adult offspring. This association is to some extent modified by offspring lifestyle factors, such as physical activity and obesity.<br>Intergenerational transmission of overweight and obesity in HUNT families | 31904500<br>29704885<br>27082110<br>25096408<br>30341129<br>27851798 |
| Gastroenterology | Greatly increased prevalence of gastro-esophageal reflux disease (GORD).<br>Improvement of GORD with weight loss and tobacco smoking cessation.<br>No increased mortality with GORD. | 22190483<br>23358462<br>24322837<br>27789657 |
| Headache | Intracranial abnormalities more common in headache sufferers<br>Migraine is not a a predictor of increased mortality<br>Previous mild head injuries: Headache more common<br>Caesarean section and the association with migraine<br>Time trends: Decreasing prevalence of migraine and MOH<br>The HUNT4 questionnaire is a valid tool to identify persons with migraine<br>Elevated CRP increased the risk of chronic migraine | 25896482<br>26115666<br>26634833<br>33208331<br>32160857<br>31195960<br>32503410 |
| Health behavior | Multiple lifestyle behaviors and mortality | 28068991 |
| Hearing | Several cardiovascular risk factors are weakly associated with hearing loss<br>Family status affects hearing loss mortality<br>Otitis media in childhood is associated with dizziness and reduced hearing in adulthood<br>Hearing loss in childhood is related to educational attainment and mental health in adulthood | 26642893<br>30463047<br>30998545<br>26335289<br>30736854<br>30946138<br>27429594 |

| | | |
|---|---|---|
| | Sociodemographic factors affect the use of hearing aids<br>The prevalence of hearing impairment has decreased in Norway the last two decades partly due to increased education, less occupational noise exposure, ear infections and smoking.<br>The use of personal music players has increased but normal use is not associated with 20-year progression in hearing. | 33509127<br>32541261<br>34181492 |
| MRI/neuroimaging | Incidental intracranial findings and their clinical impact<br>Marked effects of intracranial volume correction methods on sex differences in neuroanatomical structures<br>Perimenopausal hormone therapy is associated with regional sparing of the CA1 subfield<br>How does the accuracy of intracranial volume measurements affect normalized brain volumes? | 26950220<br>26217172<br>26130062<br>25857759 |
| Obesity | Trends in adult body-mass index in 200 countries from 1975 to 2014<br>Worldwide trends in body-mass index, underweight, overweight, and obesity from 1975 to 2016<br>Rising rural body-mass index is the main driver of the global obesity epidemic in adults<br>Social and spatial patterns of obesity diffusion over three decades<br>Central obesity is associated with lower intake of whole-grain bread, less frequent breakfast and lunch and more frequent nightly meals, when adjusted for age, sex and multiple testing. | 27115820<br>29029897<br>31068725<br>24138786<br>24833275 |
| Occupational health | Farmers' mental health: A longitudinal sibling comparison<br>Disability pension and symptoms of anxiety and depression: a prospective comparison of farmers and other occupational groups.<br>Health and unemployment: 14 years of follow-up on job loss | 27636024<br>26525724<br>26715474 |
| Oral health | Prevalence data and trends of dental caries in the Norwegian population.<br>Inequalities in dental service utilization.<br>Oral health-related quality of life in older adults.<br>Classification and current prevalence of periodontal disease among Norwegian adults. | 22779374<br>23012325<br>21226856<br>34101228 |
| Pregnancy and maternal cardiovascular health | Conventional cardiovascular risk factors are important targets for cardiovascular prevention in women with hypertensive disorders of pregnancy.<br>Adding pregnancy complications to an established CVD risk prediction model made no major improvements to CVD prediction. | 31188397<br>30596987 |

| | | |
|---|---|---|
| | Parity is associated with lasting reduction in blood pressure.<br>The association between preterm delivery and CVD is not explained by commonly measured cardiovascular risk factors. | 29980890<br><br>33022746 |
| Public health | Trends in disability-free life expectancy (DFLE) from 1995 to 2017 in the older Norwegian population<br>Employment, behavioral and psychosocial factors associated with geographical inequalities | 33908292<br><br>32861971 |
| Social epidemiology | Global effects of income and income inequality on adult height<br>Socioeconomic position, multimorbidity and mortality<br>Socioeconomic inequalities in the prevalence of complex multimorbidity | 28199042<br><br>32858852<br><br>32546494 |
| Thyroid function | Changes in the prevalence of hypothyroidism<br>Autoimmune diabetes, but not type 2 diabetes, is associated with increased prevalence of hypo- and hyperthyroidism and TPO antibodies<br>Associations of thyroid function with coronary heart disease, fractures, depressive symptoms and dementia: contributions to the Thyroid Studies Collaboration consortium | 23975540<br>26583583<br><br><br>25893284,<br>26010634,<br>28482002,<br>33154486,<br>34491268 |
