## Supplementary Table S6 for "Cohort Profile Update: The HUNT Study, Norway"

**Supplementary Table S6.** The percentage of individuals with specific ICPC-2 diagnoses recorded in the Norwegian Registry for Primary Health Care (on general practitioners' reimbursement forms to the KUHR database or in the IPLOS registry of health care services) during 2017 (for HUNT4-N, Nord-Trøndelag) or 2019 (for HUNT4-S, Sør-Trøndelag), reported separately for participants and all invitees to HUNT4-N and HUNT4-S. The numbers of people are based on residents in 2017 (for Nord-Trøndelag) or 2019 (for Sør-Trøndelag) according to the National Population Register.

| ICPC-2 diagnosis | | | Participants in HUNT4-N | | | | All residents in the Nord-Trøndelag aged 20-104 years | | | | Participants in HUNT4-S | | | | All residents in Sør-Trøndelag aged 20-104 years | | | |
|---|---|---|---|---|---|---|---|---|---|---|---|---|---|---|---|---|---|---|
| | | Sex | 20-44 years | 45-69 years | ≥70 years | **Total** | 20-44 years | 45-69 years | ≥70 years | **Total** | 20-44 years | 45-69 years | ≥70 years | **Total** | 20-44 years | 45-69 years | ≥70 years | **Total** |
| | *n* | W | 9679 | 14 335 | 6228 | **30 242** | 20 029 | 21 321 | 10 638 | **51 988** | 23 698 | 27 671 | 9304 | **60 673** | 57 675 | 46 591 | 21 381 | **125 647** |
| | *n* | M | 7124 | 12 538 | 5283 | **24 945** | 20 584 | 21 994 | 8847 | **51 425** | 15 173 | 21 959 | 8609 | **45 741** | 60 732 | 47 002 | 17 787 | **125 521** |
| D84 | Oesophagus | W | 0.5 | 1.2 | 1.9 | **1.1** | 0.6 | 1.2 | 1.9 | **1.1** | 0.6 | 1.2 | 2.0 | **1.1** | 0.6 | 1.2 | 2.3 | **1.1** |
| D84 | disease | M | 0.8 | 1.1 | 1.7 | **1.2** | 0.7 | 1.1 | 1.5 | **1.0** | 0.7 | 0.9 | 1.2 | **0.9** | 0.6 | 1.0 | 1.4 | **0.9** |
| K75 | Myocardial | W | 0.0 | 0.1 | 0.4 | **0.1** | 0.0 | 0.1 | 0.6 | **0.2** | 0.0 | 0.1 | 0.5 | **0.1** | 0.0 | 0.1 | 0.7 | **0.2** |
| K75 | infarction | M | 0.0 | 0.6 | 0.8 | **0.4** | 0.0 | 0.5 | 0.9 | **0.4** | 0.0 | 0.4 | 0.8 | **0.3** | 0.0 | 0.4 | 1.1 | **0.3** |
| K76 | Coronary artery | W | 0.0 | 0.5 | 1.6 | **0.5** | 0.0 | 0.5 | 1.9 | **0.6** | 0.0 | 0.4 | 1.7 | **0.4** | 0.0 | 0.4 | 2.4 | **0.6** |
| K76 | disease | M | 0.1 | 1.9 | 4.4 | **1.9** | 0.0 | 1.8 | 4.6 | **1.6** | 0.0 | 1.8 | 5.4 | **1.9** | 0.0 | 1.7 | 5.6 | **1.4** |
| K77 | Heart failure | W | 0.0 | 0.1 | 2.7 | **0.6** | 0.0 | 0.2 | 4.4 | **1.0** | 0.0 | 0.1 | 2.0 | **0.4** | 0.0 | 0.2 | 3.4 | **0.7** |
| K77 | | M | 0.0 | 0.2 | 2.8 | **0.7** | 0.0 | 0.4 | 4.4 | **1.0** | 0.1 | 0.3 | 2.8 | **0.7** | 0.0 | 0.4 | 4.1 | **0.7** |
| K78 | Atrial fibrillation | W | 0.0 | 0.4 | 6.8 | **1.6** | 0.0 | 0.5 | 8.3 | **1.9** | 0.0 | 0.4 | 5.8 | **1.1** | 0.0 | 0.5 | 7.3 | **1.4** |
| K78 | | M | 0.1 | 1.4 | 10.8 | **3.1** | 0.1 | 1.6 | 12.6 | **2.9** | 0.1 | 1.3 | 9.6 | **2.4** | 0.1 | 1.2 | 10.5 | **2.0** |
| K86 | Arterial hyper-. | W | 0.8 | 8.2 | 23.5 | **9.0** | 0.8 | 8.7 | 24.0 | **8.8** | 0.7 | 7.5 | 22.1 | **7.1** | 0.5 | 7.4 | 24.2 | **7.1** |
| K86 | tension, uncompl | M | 1.5 | 11.6 | 18.6 | **10.2** | 1.2 | 11.3 | 18.8 | **8.5** | 1.1 | 9.9 | 18.7 | **8.6** | 0.7 | 8.8 | 18.8 | **6.3** |
| K89 | TIA, cerebral | W | 0.1 | 0.2 | 1.5 | **0.4** | 0.1 | 0.2 | 1.7 | **0.5** | 0.0 | 0.1 | 1.0 | **0.2** | 0.0 | 0.1 | 1.4 | **0.3** |
| K89 | infarction | M | 0.1 | 0.4 | 0.9 | **0.4** | 0.0 | 0.3 | 1.2 | **0.4** | 0.0 | 0.2 | 0.9 | **0.3** | 0.0 | 0.2 | 1.0 | **0.2** |
| K90 | Stroke | W | 0.1 | 0.4 | 2.6 | **0.8** | 0.0 | 0.6 | 3.7 | **1.0** | 0.0 | 0.2 | 2.5 | **0.5** | 0.0 | 0.4 | 3.7 | **0.8** |
| K90 | | M | 0.1 | 0.7 | 3.2 | **1.1** | 0.1 | 0.9 | 4.4 | **1.2** | 0.0 | 0.6 | 2.4 | **0.7** | 0.0 | 0.7 | 3.9 | **0.8** |
| L18 | Myalgia | W | 3.7 | 5.4 | 3.0 | **4.4** | 3.8 | 5.7 | 2.8 | **4.4** | 2.5 | 3.6 | 2.3 | **3.0** | 2.3 | 3.9 | 2.3 | **2.9** |
| L18 | | M | 1.4 | 1.7 | 1.0 | **1.5** | 1.4 | 1.8 | 1.0 | **1.5** | 0.7 | 1.0 | 0.9 | **0.9** | 0.8 | 1.0 | 0.8 | **0.9** |
| L83 | Cervical | W | 0.7 | 1.5 | 0.9 | **1.2** | 0.8 | 1.6 | 0.8 | **1.1** | 0.5 | 1.0 | 0.5 | **0.8** | 0.5 | 1.0 | 0.5 | **0.7** |
| L83 | spondylosis | M | 0.5 | 1.3 | 0.5 | **0.9** | 0.5 | 1.4 | 0.5 | **0.9** | 0.3 | 0.8 | 0.4 | **0.5** | 0.3 | 0.8 | 0.4 | **0.5** |

| | | | | | | | | | | | | | | | | | | |
|---|---|---|---|---|---|---|---|---|---|---|---|---|---|---|---|---|---|---|
| L84 | Lumbal pain | W | 1.5 | 1.8 | 2.4 | **1.8** | 1.3 | 1.8 | 2.4 | **1.7** | 1.1 | 1.3 | 2.3 | **1.4** | 1.0 | 1.5 | 2.4 | **1.4** |
| L84 | | M | 1.7 | 1.9 | 1.9 | **1.8** | 1.7 | 2.1 | 1.8 | **1.9** | 1.1 | 1.4 | 1.7 | **1.4** | 1.1 | 1.5 | 1.7 | **1.3** |
| L88 | Rheumatois | W | 0.6 | 1.8 | 2.6 | **1.6** | 0.6 | 2.0 | 2.7 | **1.6** | 0.7 | 1.5 | 2.6 | **1.4** | 0.5 | 1.4 | 2.8 | **1.3** |
| L88 | arthritis | M | 0.4 | 1.0 | 1.3 | **0.9** | 0.3 | 1.1 | 1.5 | **0.8** | 0.4 | 1.0 | 1.2 | **0.9** | 0.3 | 0.9 | 1.3 | **0.6** |
| L95 | Osteoporosis | W | 0.0 | 1.1 | 5.4 | **1.7** | 0.0 | 1.2 | 5.9 | **1.7** | 0.0 | 1.5 | 6.5 | **1.7** | 0.0 | 1.5 | 7.6 | **1.9** |
| L95 | | M | 0.0 | 0.1 | 0.5 | **0.2** | 0.0 | 0.2 | 0.7 | **0.2** | 0.0 | 0.2 | 0.9 | **0.3** | 0.0 | 0.2 | 1.0 | **0.2** |
| N01 | Headache | W | 3.7 | 2.1 | 1.4 | **2.5** | 3.6 | 2.0 | 1.4 | **2.5** | 3.4 | 2.2 | 1.7 | **2.6** | 3.4 | 2.3 | 1.7 | **2.7** |
| N01 | | M | 1.5 | 1.3 | 1.2 | **1.3** | 1.6 | 1.3 | 1.2 | **1.4** | 1.4 | 1.3 | 1.1 | **1.3** | 1.6 | 1.3 | 1.0 | **1.4** |
| N17 | Dizziness/ | W | 2.5 | 2.5 | 5.1 | **3.0** | 2.3 | 2.4 | 4.7 | **2.8** | 2.4 | 2.6 | 5.7 | **3.0** | 2.2 | 2.7 | 6.0 | **3.1** |
| N17 | vertigo | M | 0.9 | 1.4 | 2.8 | **1.6** | 0.8 | 1.3 | 3.0 | **1.4** | 1.0 | 1.4 | 3.5 | **1.7** | 0.9 | 1.4 | 3.7 | **1.5** |
| N89 | Migraine | W | 2.4 | 1.1 | 0.2 | **1.3** | 2.4 | 1.1 | 0.2 | **1.4** | 3.0 | 1.6 | 0.4 | **1.9** | 2.7 | 1.6 | 0.4 | **1.9** |
| N89 | | M | 0.7 | 0.4 | 0.1 | **0.4** | 0.7 | 0.4 | 0.1 | **0.4** | 0.8 | 0.3 | 0.2 | **0.4** | 0.7 | 0.4 | 0.1 | **0.5** |
| P06 | Insomnia | W | 2.1 | 2.5 | 3.7 | **2.7** | 2.1 | 2.8 | 3.8 | **2.7** | 2.5 | 3.2 | 3.0 | **2.9** | 2.6 | 3.4 | 3.8 | **3.1** |
| P06 | | M | 1.8 | 1.6 | 2.1 | **1.8** | 1.9 | 1.9 | 2.3 | **1.9** | 2.0 | 2.2 | 2.4 | **2.1** | 2.1 | 2.3 | 2.4 | **2.2** |
| P15 | Alcohol abuse | W | 0.0 | 0.2 | 0.1 | **0.1** | 0.1 | 0.4 | 0.2 | **0.2** | 0.1 | 0.2 | 0.1 | **0.1** | 0.1 | 0.4 | 0.3 | **0.3** |
| P15 | | M | 0.1 | 0.5 | 0.3 | **0.4** | 0.3 | 1.0 | 0.4 | **0.6** | 0.3 | 0.5 | 0.5 | **0.4** | 0.4 | 0.9 | 0.8 | **0.6** |
| P20 | Memory | W | 0.4 | 0.4 | 1.8 | **0.7** | 0.4 | 0.4 | 2.2 | **0.8** | 0.4 | 0.3 | 1.5 | **0.5** | 0.4 | 0.5 | 2.8 | **0.8** |
| P20 | distubancis | M | 0.2 | 0.3 | 1.5 | **0.6** | 0.2 | 0.3 | 2.1 | **0.6** | 0.3 | 0.3 | 1.3 | **0.5** | 0.3 | 0.4 | 2.4 | **0.6** |
| P70 | Dementia | W | 0.0 | 0.1 | 2.6 | **0.6** | 0.0 | 0.2 | 5.7 | **1.2** | 0.0 | 0.0 | 3.1 | **0.5** | 0.0 | 0.1 | 6.8 | **1.2** |
| P70 | | M | 0.0 | 0.1 | 1.7 | **0.4** | 0.0 | 0.1 | 3.8 | **0.7** | 0.0 | 0.1 | 1.8 | **0.4** | 0.0 | 0.1 | 4.4 | **0.7** |
| P74 | Anxiety | W | 1.6 | 1.1 | 1.3 | **1.3** | 1.7 | 1.5 | 1.9 | **1.6** | 1.7 | 1.0 | 0.9 | **1.2** | 1.8 | 1.4 | 2.0 | **1.7** |
| P74 | | M | 0.7 | 0.6 | 0.4 | **0.6** | 0.9 | 0.9 | 0.7 | **0.8** | 1.1 | 0.6 | 0.4 | **0.7** | 1.0 | 0.9 | 0.6 | **0.9** |
| P76 | Depression | W | 4.8 | 3.8 | 3.5 | **4.1** | 5.3 | 4.5 | 4.5 | **4.8** | 4.8 | 2.7 | 2.2 | **3.5** | 4.8 | 3.4 | 3.3 | **4.0** |
| P76 | | M | 2.8 | 2.1 | 1.3 | **2.1** | 3.2 | 2.5 | 1.8 | **2.7** | 2.9 | 1.8 | 1.2 | **2.0** | 2.7 | 2.1 | 1.6 | **2.3** |
| R75 | Acute sinusitis | W | 4.0 | 3.0 | 1.3 | **3.0** | 3.8 | 2.9 | 1.0 | **2.8** | 2.9 | 2.4 | 1.1 | **2.4** | 2.7 | 2.3 | 0.9 | **2.2** |
| R75 | | M | 1.8 | 1.6 | 1.2 | **1.5** | 1.6 | 1.4 | 0.8 | **1.4** | 1.3 | 1.2 | 0.7 | **1.1** | 1.1 | 1.0 | 0.6 | **1.0** |
| R95 | COPD | W | 0.1 | 1.3 | 4.5 | **1.6** | 0.0 | 1.8 | 5.6 | **1.9** | 0.0 | 1.0 | 3.3 | **1.0** | 0.0 | 1.4 | 5.2 | **1.4** |
| R95 | | M | 0.1 | 1.0 | 5.2 | **1.6** | 0.0 | 1.4 | 6.5 | **1.7** | 0.0 | 0.9 | 4.1 | **1.2** | 0.0 | 1.2 | 5.6 | **1.3** |
| R96 | Asthma | W | 1.7 | 1.9 | 2.3 | **1.9** | 1.6 | 2.1 | 2.4 | **2.0** | 1.3 | 1.7 | 2.3 | **1.6** | 1.3 | 1.7 | 2.5 | **1.6** |
| R96 | | M | 1.4 | 1.7 | 1.6 | **1.6** | 1.2 | 1.7 | 1.5 | **1.5** | 1.1 | 1.3 | 1.4 | **1.2** | 0.9 | 1.2 | 1.4 | **1.1** |
| R97 | Allergic rhinitis | W | 2.5 | 1.4 | 0.6 | **1.6** | 2.3 | 1.4 | 0.6 | **1.6** | 2.1 | 1.1 | 0.7 | **1.4** | 2.1 | 1.0 | 0.6 | **1.4** |

| | | | | | | | | | | | | | | | | | | |
|---|---|---|---|---|---|---|---|---|---|---|---|---|---|---|---|---|---|---|
| R97 | | M | 2.3 | 1.0 | 0.8 | **1.3** | 1.8 | 1.0 | 0.7 | **1.3** | 1.8 | 0.7 | 0.5 | **1.0** | 1.7 | 0.7 | 0.5 | **1.1** |
| S91 | Psoriasis | W | 0.4 | 1.0 | 0.8 | **0.8** | 0.4 | 1.0 | 1.1 | **0.8** | 0.5 | 0.9 | 0.9 | **0.7** | 0.4 | 0.9 | 1.1 | **0.7** |
| S91 | | M | 0.6 | 0.9 | 0.8 | **0.8** | 0.4 | 0.9 | 0.8 | **0.7** | 0.5 | 0.7 | 0.7 | **0.6** | 0.4 | 0.7 | 0.9 | **0.6** |
| T82 | Obesity | W | 0.4 | 0.4 | 0.2 | **0.3** | 0.4 | 0.4 | 0.1 | **0.4** | 0.4 | 0.3 | 0.1 | **0.3** | 0.3 | 0.3 | 0.1 | **0.3** |
| T82 | | M | 0.1 | 0.2 | 0.1 | **0.1** | 0.2 | 0.2 | 0.1 | **0.2** | 0.2 | 0.3 | 0.1 | **0.2** | 0.1 | 0.3 | 0.1 | **0.2** |
| T83 | Overweight | W | 1.0 | 0.9 | 0.3 | **0.8** | 1.1 | 1.1 | 0.3 | **0.9** | 1.2 | 1.3 | 0.3 | **1.1** | 0.9 | 1.3 | 0.3 | **0.9** |
| T83 | | M | 0.5 | 0.7 | 0.2 | **0.5** | 0.5 | 0.8 | 0.2 | **0.6** | 0.5 | 0.6 | 0.2 | **0.5** | 0.4 | 0.7 | 0.2 | **0.5** |
| T85 | Hyperthyroidism | W | 0.2 | 0.5 | 0.4 | **0.4** | 0.3 | 0.5 | 0.5 | **0.4** | 0.3 | 0.3 | 0.4 | **0.3** | 0.3 | 0.4 | 0.6 | **0.4** |
| T85 | | M | 0.1 | 0.1 | 0.1 | **0.1** | 0.0 | 0.1 | 0.2 | **0.1** | 0.1 | 0.1 | 0.1 | **0.1** | 0.0 | 0.1 | 0.1 | **0.1** |
| T86 | Hypothyroidism | W | 1.5 | 3.7 | 4.9 | **3.2** | 1.4 | 3.5 | 5.4 | **3.1** | 1.7 | 3.8 | 5.6 | **3.3** | 1.5 | 3.5 | 6.0 | **3.0** |
| T86 | | M | 0.3 | 0.8 | 1.5 | **0.8** | 0.2 | 0.8 | 1.7 | **0.7** | 0.3 | 0.7 | 1.5 | **0.7** | 0.2 | 0.7 | 1.7 | **0.6** |
| T89 | Diabetes type 1 | W | 0.1 | 0.3 | 0.5 | **0.3** | 0.2 | 0.4 | 1.1 | **0.5** | 0.3 | 0.3 | 0.7 | **0.3** | 0.2 | 0.4 | 1.0 | **0.4** |
| T89 | | M | 0.2 | 0.5 | 0.9 | **0.5** | 0.2 | 0.6 | 1.3 | **0.6** | 0.3 | 0.6 | 1.0 | **0.6** | 0.3 | 0.7 | 1.4 | **0.6** |
| T90 | Diabetes type 2 | W | 0.4 | 3.2 | 8.3 | **3.4** | 0.5 | 4.0 | 9.6 | **3.8** | 0.3 | 2.7 | 6.8 | **2.4** | 0.4 | 3.2 | 8.5 | **2.8** |
| T90 | | M | 0.5 | 5.8 | 11.0 | **5.4** | 0.6 | 6.4 | 12.2 | **5.1** | 0.5 | 4.9 | 10.0 | **4.4** | 0.5 | 5.2 | 11.3 | **3.8** |
| T92 | Arthritis | W | 0.1 | 0.2 | 1.1 | **0.4** | 0.1 | 0.3 | 1.4 | **0.4** | 0.0 | 0.2 | 0.9 | **0.2** | 0.0 | 0.2 | 1.2 | **0.3** |
| T92 | hyperurecemia | M | 0.4 | 1.1 | 2.8 | **1.2** | 0.3 | 1.1 | 2.7 | **1.0** | 0.3 | 1.0 | 2.6 | **1.1** | 0.2 | 1.0 | 2.7 | **0.9** |
| U04 | Urinary | W | 0.5 | 1.0 | 2.7 | **1.2** | 0.5 | 1.0 | 3.0 | **1.2** | 0.4 | 0.7 | 2.9 | **0.9** | 0.3 | 0.8 | 3.4 | **1.0** |
| U04 | incontinence | M | 0.1 | 0.3 | 1.3 | **0.5** | 0.1 | 0.3 | 1.4 | **0.4** | 0.1 | 0.2 | 0.9 | **0.3** | 0.0 | 0.3 | 1.5 | **0.3** |

Abbreviations: COPD, chronic obstructive pulmonary disease; ICPC, International Classification of Primary Care; IPLOS, Individbasert pleie- og omsorgsstatistikk (Individual-based nursing and care statistics); KUHR, Kontroll og utbetaling av helserefusjoner (Control and payment of health reimbursements)
