## Supplementary Table S7 for "Cohort Profile Update: The HUNT Study, Norway"

**Supplementary Table S7.** Validity of self-reported conditions among participants of HUNT4-N, using as reference standards diagnoses recorded in the local/regional hospitals (ICD-9 or 10, 1987-2019) or general practitioner diagnoses sent to the KUHR database for reimbursement purposes (ICPC-2, 2006-2019).

| Self-reported diseases and conditions | ICPC-2 | ICD-9 | ICD-10 | Sensitivity, % | Specificity, % | Positive predictive value, % | Negative predictive value, % |
|---|---|---|---|---|---|---|---|
| Do you have, or have you ever had | | | | | | | |
| Angina | K74 | 413 | I20 | 34 | 99.3 | 78 | 95.3 |
| Myocardial infarction | K75 | 410, 411, 412 | I21, I24, I25.2 | 76 | 99.6 | 91 | 98.9 |
| Heart failure | K77 | 425, 428 | I42, I50 | 39 | 99.2 | 56 | 98.4 |
| Atrial fibrillation | K78 | 427.3 | I48 | 69 | 97.9 | 63 | 98.4 |
| Stroke | K90, K91 | 430, 431, 432, 433, 434 | I60, I61, I63, I64 | 59 | 99.1 | 76 | 98.1 |
| Asthma | R96 | 493 | J45, J46 | 66 | 95.6 | 68 | 95.1 |
| COPD or emphysema | R95 | 491, 492 | J43, J44 | 48 | 99.5 | 84 | 97.2 |
| Diabetes | T89, T90 | | | 81 | 99.5 | 92 | 98.6 |
| Hypo- or hyperthyroidism | T85, T86 | | | 84 | 98.2 | 79 | 98.7 |
| Migraine | N89 | | | 84 | 89 | 34 | 98.8 |
| Psoriasis | S91 | | | 72 | 96 | 46 | 98.7 |
| Rheumatoid arthritis | | 714 | M05, M06, M08 | 63 | 95.9 | 27 | 99.1 |
| Ankylosing spondylitis | | 720 | M45 | 53 | 99.4 | 63 | 99.1 |
| Gout | T92 | | | 73 | 98.3 | 60 | 99.1 |
| Mental problems you have sought help for | P01, P02, P03, P74, P76 | | | 41 | 95 | 79 | 76 |
| Allergic rhinitis | R75, R97 | | | 47 | 81 | 52 | 78 |
| Much heartburn last 12 months | D84 | | | 74 | 63 | 15 | 96.5 |
| Current use of antihypertensive medication | K85, K86, K87 | | | 75 | 96 | 88 | 91 |
| Ongoing musculoskeletal pain for at least 3 months during the last 12 months | L18, L83, L84, L86 | | | 68 | 56 | 52 | 71 |
| Headache during the last 12 months | N01, N89 | | | 68 | 70 | 34 | 90 |
| Sought GP for urinary incontinence | U04 | | | 74 | 93 | 44 | 97.9 |

Abbreviations: COPD, chronic obstructive pulmonary disease; GP, general practitioner; ICD, International Classification of Diseases; ICPC, International Classification of Primary Care; KUHR, Kontroll og utbetaling av helserefusjoner (Control and payment of health reimbursements)
