## Supplementary Figure S1 for "Cohort Profile Update: The HUNT Study, Norway"

**Supplementary Figure S1.** Primary health care utilization (a. number of general practitioner visits, b. percentage using home nursing, c. percentage staying in retirement/nursing homes) among participants and nonparticipants of HUNT4-N (in 2017) and HUNT4-S (in 2019), separately among women (left panels) and men (right panels), according to the Norwegian Registry for Primary Health Care.

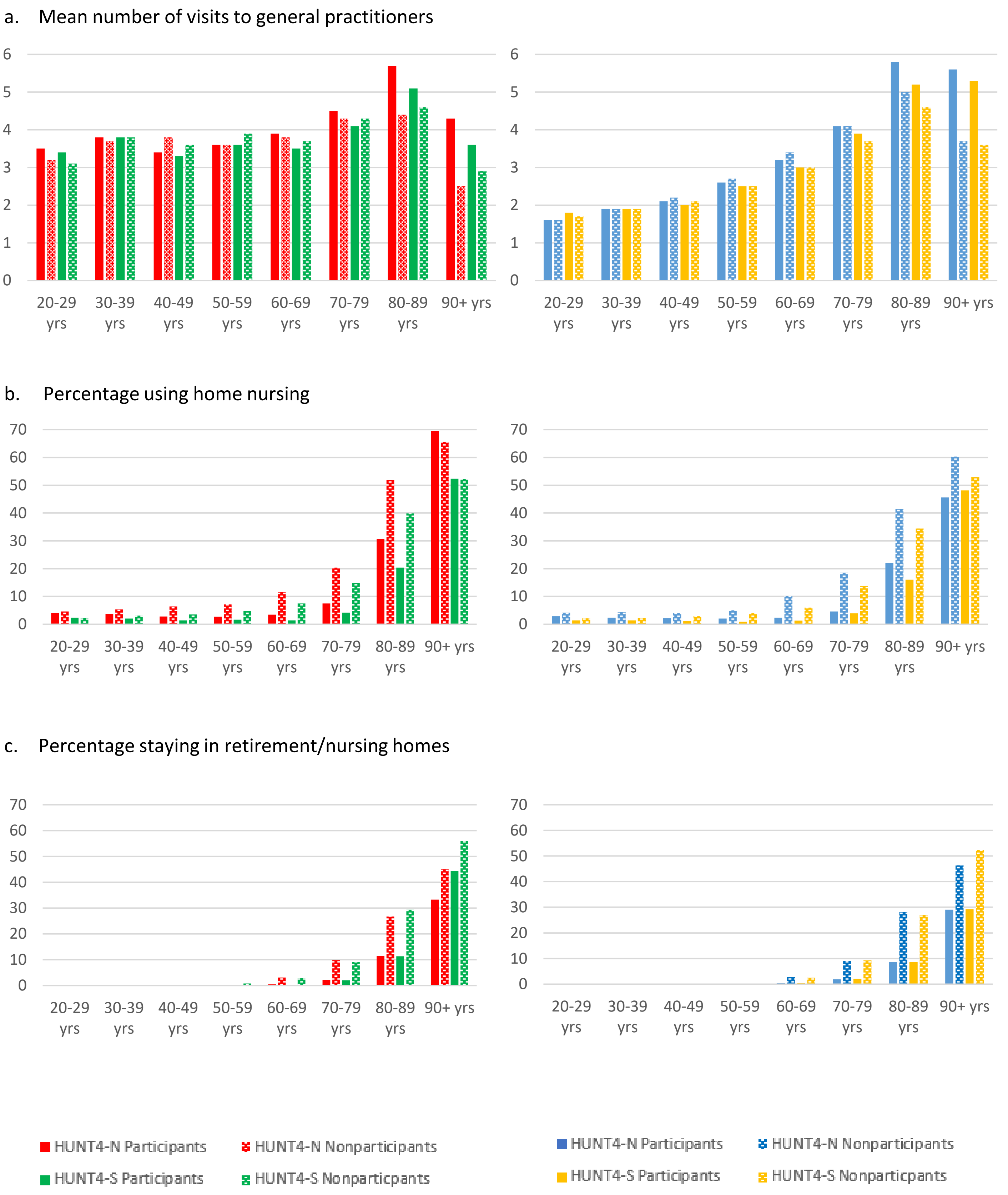
